# The landscape and level of alcohol policy enforcement in Tanzania

**DOI:** 10.1101/2024.04.17.24305975

**Authors:** Kim Madundo, Aliza Hudda, Maaike L. Seekles, Blandina Mmbaga, Angela Obasi

**Affiliations:** Department of Mental Health and Psychiatry, Kilimanjaro Christian Medical University College, Moshi, Tanzania; Department of Mental Health and Psychiatry, Kilimanjaro Christian Medical Centre, Moshi, Tanzania; The Liverpool School of Tropical Medicine, Department of International Public Health, Liverpool, UK; Department of Paediatric and Child Health, Kilimanjaro Christian Medical University College, Moshi, Tanzania; Kilimanjaro Clinical Research Institute, Moshi, Tanzania; AXESS Sexual Health, Royal Liverpool University Hospitals NHS Foundation Trust, Liverpool, UK

## Abstract

Harmful use of alcohol causes more deaths in Sub-Saharan Africa than in any other region. In Tanzania, where alcohol use disorders rates are twice the overall African average, harmful alcohol consumption is a public health concern. Given the lack of a contemporary overview of the alcohol policy landscape, we conducted a mixed-methods review of key alcohol-related policies, implementers, and initiatives in Tanzania. We conducted a desk-based review of policy-related documents, and in-depth interviews with eight key informants guided by the 10-composite-indicator framework of a tool for measuring alcohol policy implementation developed by World Health Organization. Representatives were from health-service delivery, community-based organizations, governmental organizations, research, and policymakers whose work is related to alcohol in Tanzania. Data was collected in October 2021, June 2022, and finalized in March 2023. Findings were analyzed using Microsoft Word v2021. Themes were identified, collected, combined, and tabulated. Differences were then resolved by first and second authors. Our findings revealed no single comprehensive national alcohol policy. Pending finalization of a draft policy, various documents and actors govern alcohol production, distribution, licensing, and consumption. Little intersectoral linkage between entities contributes to poor enforcement of these regulations. Regulation is stronger in urban areas, and restrictions more effective on industrial alcohol. However, the majority of consumed alcohol in Tanzania is informally-produced, especially in rural settings. Socio-cultural context plays a key role in alcohol production and consumption, contributing to early-age exposure to alcohol. Alcohol is a growing source of revenue for the Tanzanian government and, therefore, imposing further restrictions is a low priority. There are important policy gaps in various sectors pertaining to alcohol regulation. Our results strongly suggest the need for a comprehensive approach to developing an overarching alcohol policy, with involvement of key stakeholders, stronger enforcement, and increased awareness, resources, and collaborations.

## Introduction

The harmful use of alcohol causes approximately 3 million deaths every year worldwide (1). In Sub-Saharan Africa (SSA), alcohol accounts for more deaths than in any other region. The reduction of harmful alcohol use is a particular priority in Tanzania, where alcohol use disorders are twice, and rates of heavy episodic drinking are six times, the Africa Region average (2). Much of this increased risk begins in adolescence (10-19y). Early (aged <15y) alcohol initiation is common, and is more likely to cause dependency issues compared to those who begin drinking above 21 (2). Adults with violence-related injuries are more than six times as likely to have a history of harmful alcohol use (3). Hazardous alcohol consumption is associated with higher odds of a comorbid mental disorder than in the general population (4). Harmful drinking has been linked to insufficient regulations and restrictions on exposure to alcohol-related cues (2,5). There is a need to delay and decrease alcohol use, but a lack of national-level integrated regulations to reinforce this (6). Our study aimed to gain an understanding of alcohol related policy and legislation, and to understand how the enforcement of such policies may affect alcohol use.

Published by WHO in 2018, The *Global Strategy to reduce the harmful use of alcohol* is the most comprehensive international alcohol policy document. This report is endorsed by World Health Organization (WHO) Member States, who commit to periodically provide country-level data on alcohol consumption and related harm, and develop and implement national alcohol policies. The strategy recommends a portfolio of national-level policy options and interventions across ten target areas that could be considered for implementation: Leadership, awareness, and commitment; health services’ response; community action; drink-driving policies and countermeasures; availability of alcohol; marketing of alcoholic beverages; pricing policies; reducing the negative consequences of drinking and alcohol intoxication; reducing the public health impact of illicit alcohol and informally produced alcohol; and monitoring and surveillance (2). Through these standard ten markers, this framework serves as a reference point for measurement and comparison of enforcement and implementation across countries worldwide. An action plan (2022–2030) was recently adopted to guide the effective implementation of the strategy (7). Separately, in 2019, the East African Community (EAC) regional policy intended to comprehensively address disparity in policies, programs, and practices, and reduce negative outcomes related to harmful use of alcohol and other drugs (8).

Aside from a page in the Global Status Report on Alcohol and Health (2018), there is no recent and detailed overview of what the alcohol policy landscape in Tanzania looks like. In response, this paper presents the findings of an iterative, mixed-methods policy review. It aims to provide an overview of key alcohol-related policies and enforced alcohol control measures, including national-level initiatives and stakeholders.

## Materials and Methods

Our review was conducted in two stages: a desk-based review of policy-related documents in Tanzania, and in-depth interviews (IDIs) with key informants. WHO-recommended target areas and corresponding indicators for alcohol-related policy and interventions provided the framework for the searches and IDIs and analysis. National-level key informants, initiatives, and implementers who could offer insight into alcohol-related policies and their enforcement across Tanzania were purposively selected.

### Data collection

Firstly, two researchers conducted a desk-based review starting on 17th October 2022, which was then updated in March 2023 to ensure any more recent documents were included. A broad Google Search was conducted, using the following terms: ‘[Policy OR Law OR Act OR regulation OR legislation] AND [Alcohol OR Liquor OR Brew] AND [Tanzania]’. This returned 80 unique and relevant results.

This open search was followed by targeted searches, informed by WHO target areas and indicators for policy implementation (9,10) and a landscape analysis of alcohol-related policies in Tanzania published in 2012 (11).

Relevant search results were downloaded to a shared folder. Content eligible for inclusion was that describing key policy/actors/plans/strategies related to alcohol consumption in Tanzania, and whereby majority of the content was related to alcohol consumption, and enforced or applicable in Tanzania. Information related to policies and interventions was extracted and a directory of all Tanzanian ministries and special offices with governance over alcohol-related activities was created (*Figure 1*). Websites of these actors were searched for any final relevant sources.

**Figure 1.**
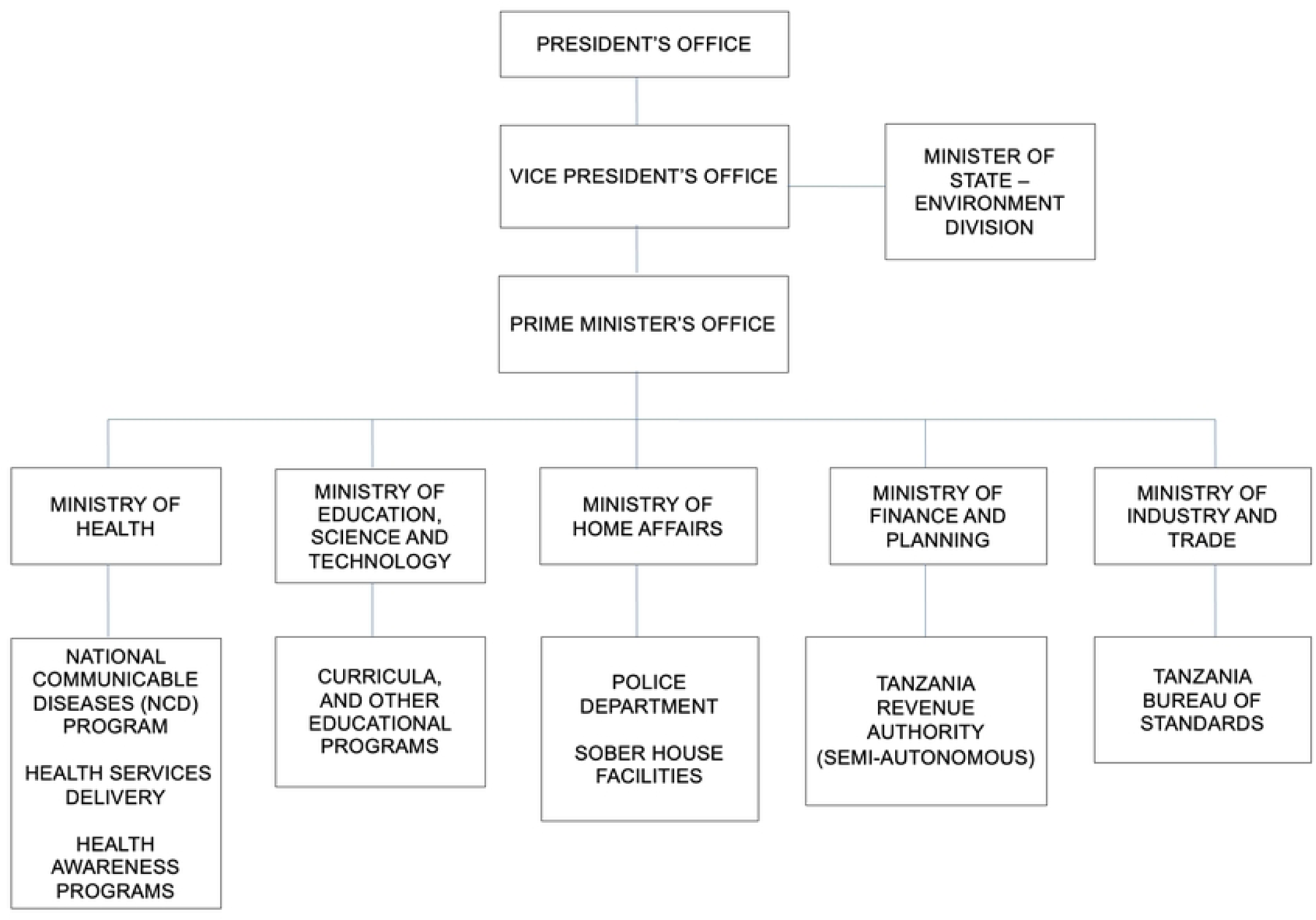
Directory of government actors essential to alcohol-related policies in Tanzania.

Secondly, six IDIs were conducted beginning from 10^th^ June 2022, and another two ending on 31^st^ March 2023. In addition to perspectives from relevant stakeholders, interviews gathered further information on policies and initiatives related to WHO target areas. Informants were representatives of health-service delivery, community-based organizations (CBOs), governmental organizations, researchers, and policymakers whose work is related to alcohol consumption in Tanzania (see Table 1).

**Table 1.** Participant characteristics.

| Participant Number | Gender | Years of Work Experience | Private/Public Sector | Scale/Level of Operations |
| --- | --- | --- | --- | --- |
| #1 | Male | 19 | Private | Regional |
| #2 | Male | 12 | Public | Regional |
| #3 | Male | 20 | Private | National |
| #4 | Male | 20 | Private | Regional |
| #5 | Male | 10 | Public | National |
| #6 | Male | 5 | Public | Regional |
| #7 | Male | 7 | Public | National |
| #8 | Female | 4 | Private | City |

We decided on this number of IDIs based on the overlap in responders’ areas of expertise and having reached saturation across our data collection. Topic guide questions were designed to cover all target areas (e.g., relevant policies, health service delivery, efforts to combat harmful alcohol use in the community, schools, and workplace, and availability of alcohol), but were adapted to fit with participant’s area of expertise/work (Table 2).

**Table 2.**
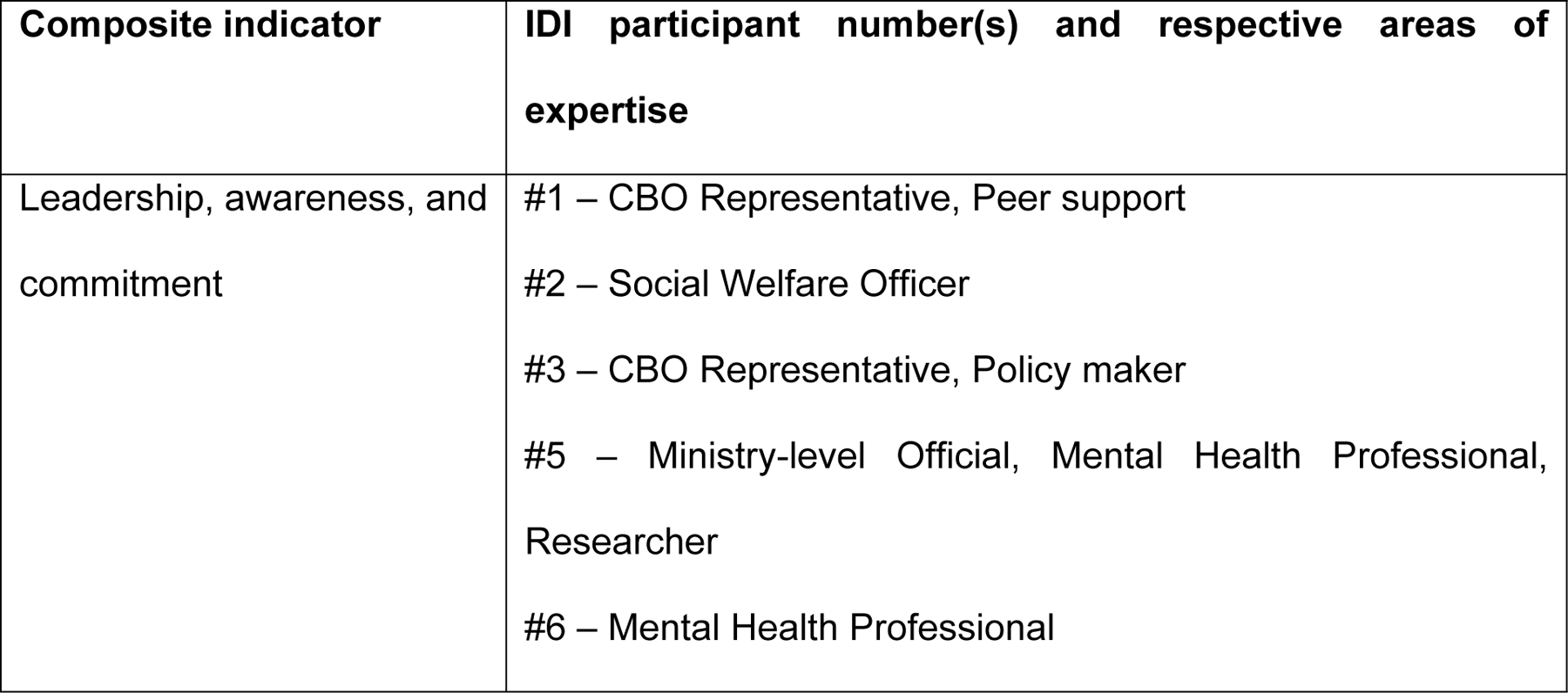
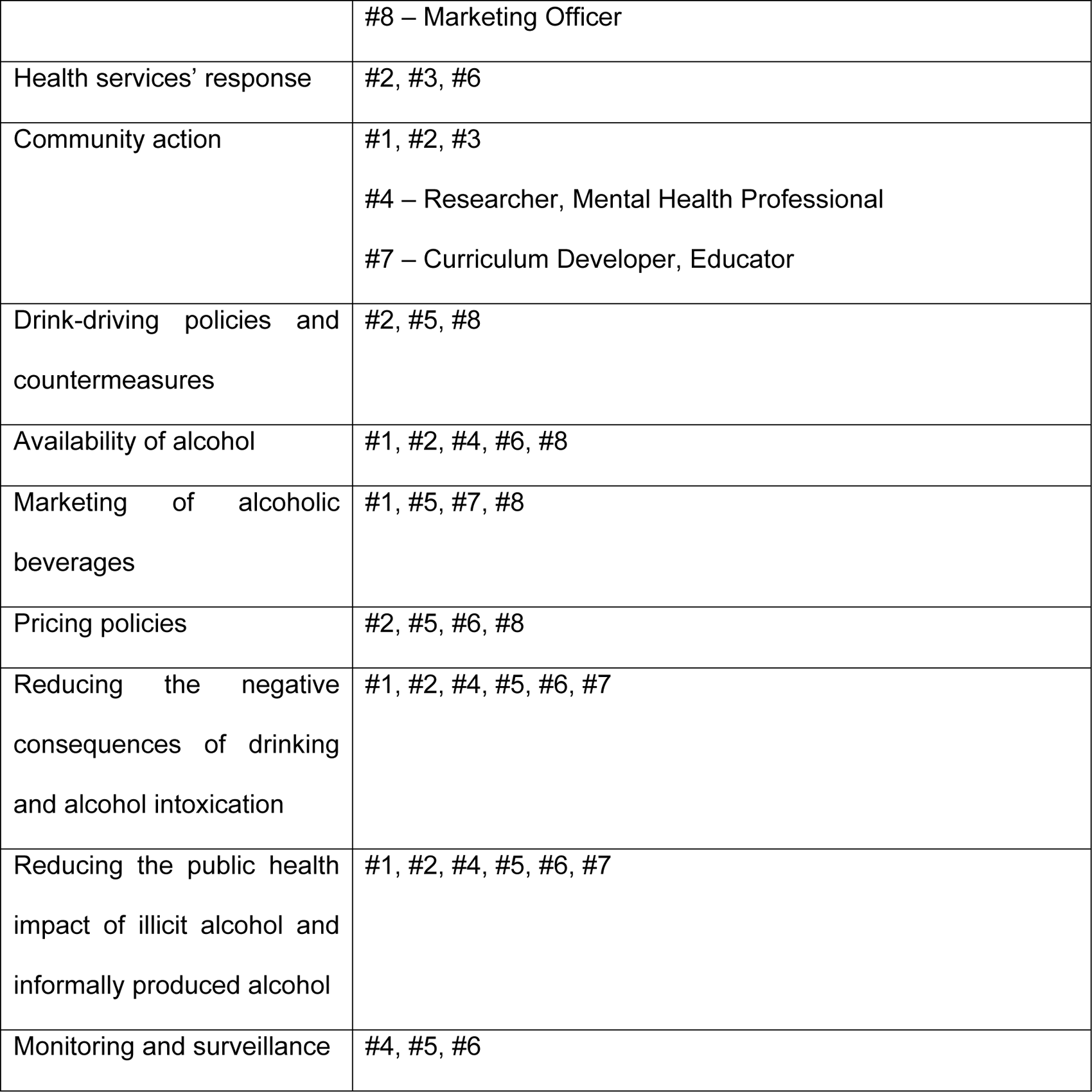
In-depth interview content, participants interviewed and their respective areas of expertise.

IDIs lasted an average of 40 minutes, were conducted in English or Kiswahili, and either in-person or virtually through video communications platform ‘Zoom’ depending on location and preference of informant. Each interview was given a code number. Participants were reimbursed 10,000 Tanzanian shillings for their participation. No identifying information was collected or recorded during IDIs.

IDI recordings were transferred from voice-recorder to a password-protected computer only accessed by KM and AH, who also transcribed the IDIs. These were then translated to English by the research team, allowing for triangulation of the translations for accuracy. Once complete, transcriptions, translations, and IDI recordings were uploaded to a secure server.

### Analysis

Google search results were analyzed by KM and AH; themes were identified, collected and combined in tabulated form. Differences were then resolved by KM and AH, and MS was consulted upon any further disagreements.

IDIs were analyzed using Microsoft Word v2021. Quotes were interpreted, color-coded and grouped according to their thematic areas based on WHO target areas, by the first author. A checklist was created indicating the status of policy items in Tanzania; absent, partial restriction, or total restriction.

To ensure trustworthiness of our findings we used interviewer triangulation; two researchers worked separately, then discussed and resolved differences. A third researcher monitored quality control of transcripts and codes. All data were triangulated and brought together in one Table, to provide an overview of the current alcohol policy landscape in Tanzania.

### Ethical approval and consent to participate

Clearance to conduct this study was granted by research committees at Kilimanjaro Christian Medical University College (Ref. #2551), the National Institute for Medical Research (Ref: NIMR/HQ/R.8a/Vol.IX/3932, and Liverpool School of Tropical Medicine (Ref: 21-075). Prior to IDIs, informants were sent an information sheet describing the purpose of the study and written informed consent was obtained.

## Results

Findings from our online searches and IDIs are presented according to the framework of composite indicators provided by WHO. See Table 3 for an overview of the reviewed policy-related documents, along with their linking components within the WHO framework.

**Table 3.** Relevant policy-related documents Error! Reference source not found.

**Table 4.**
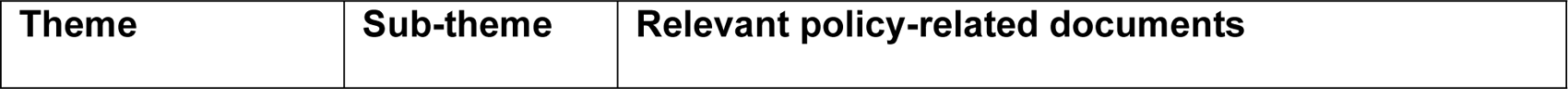
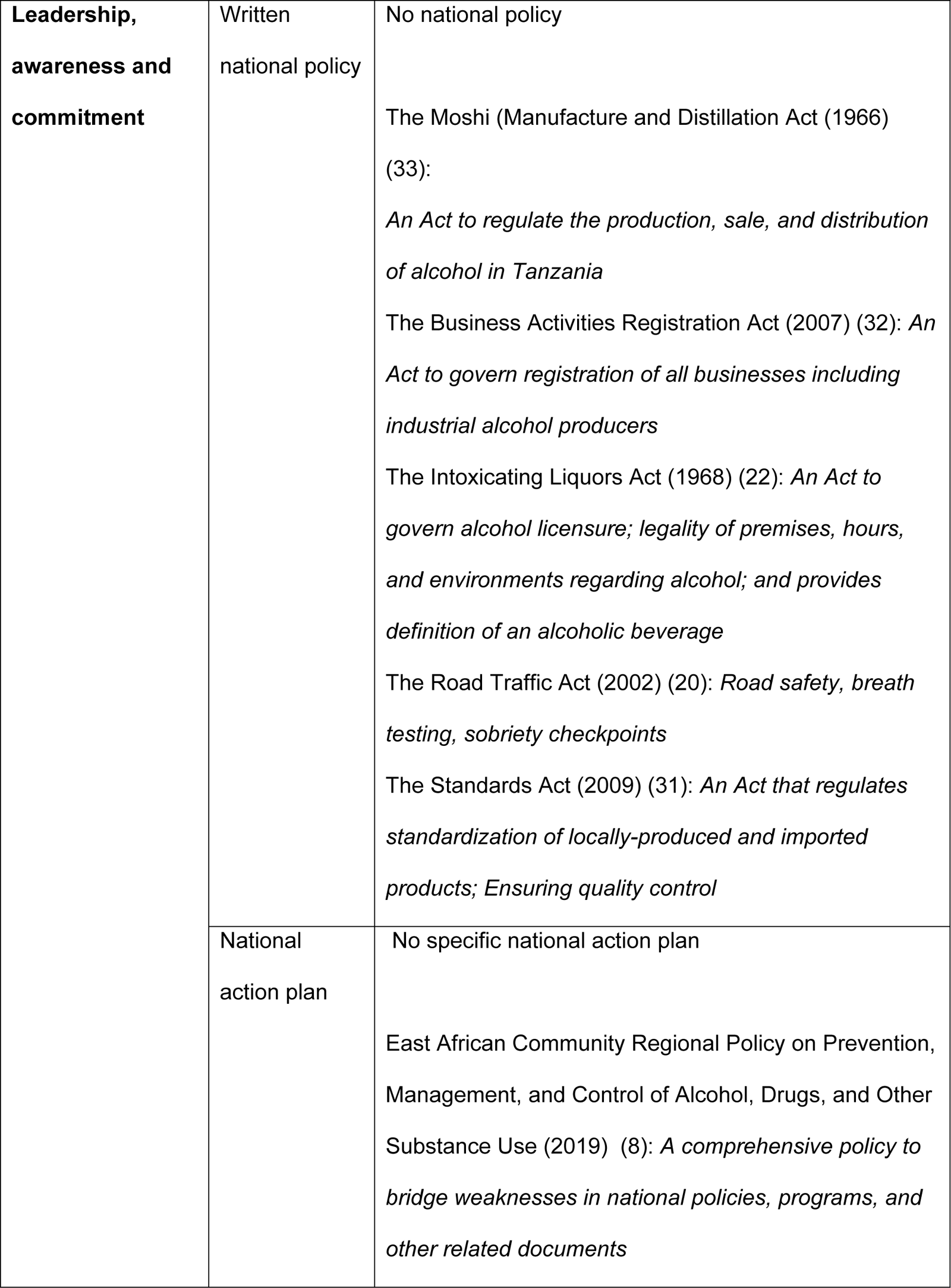
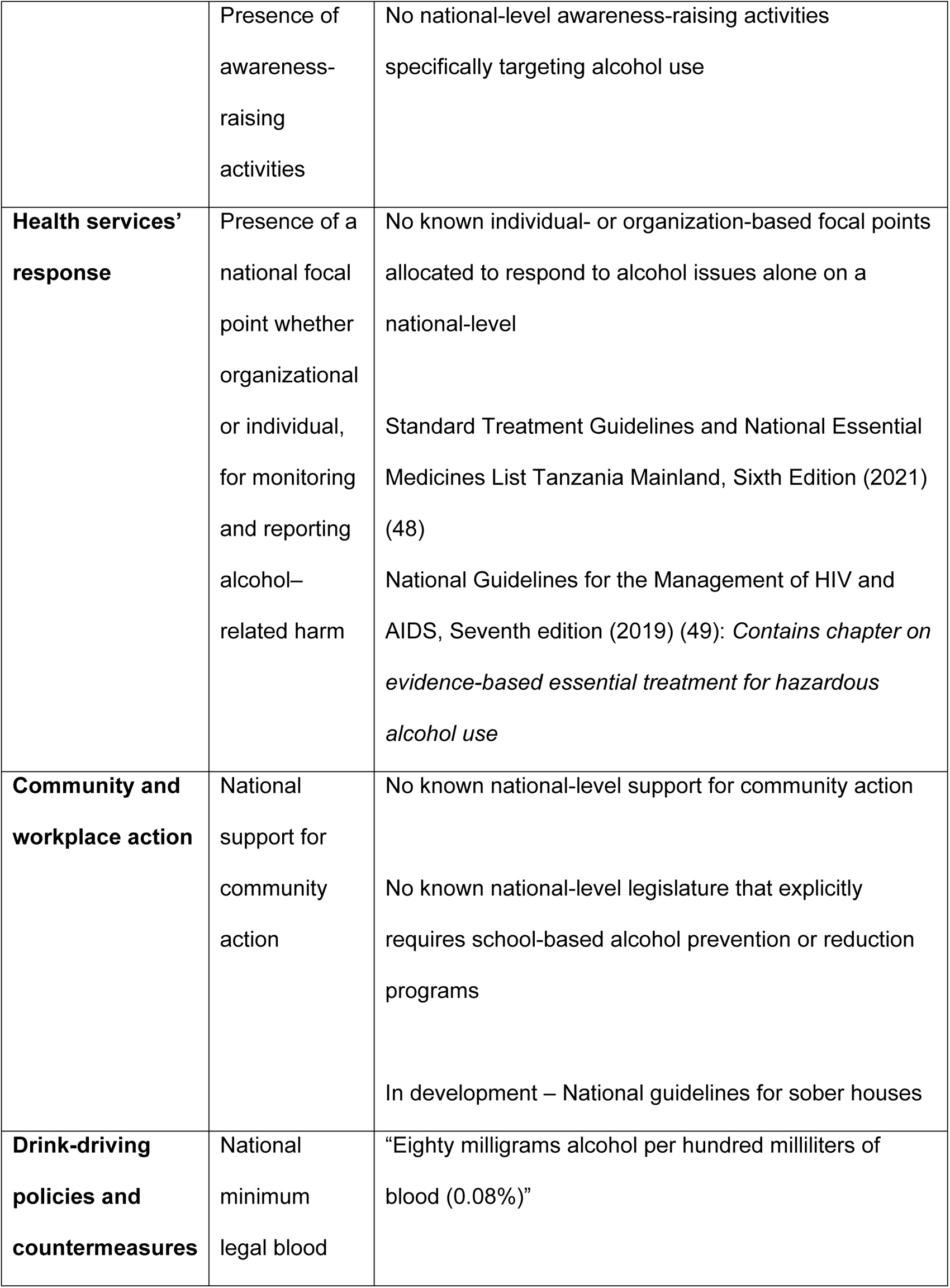
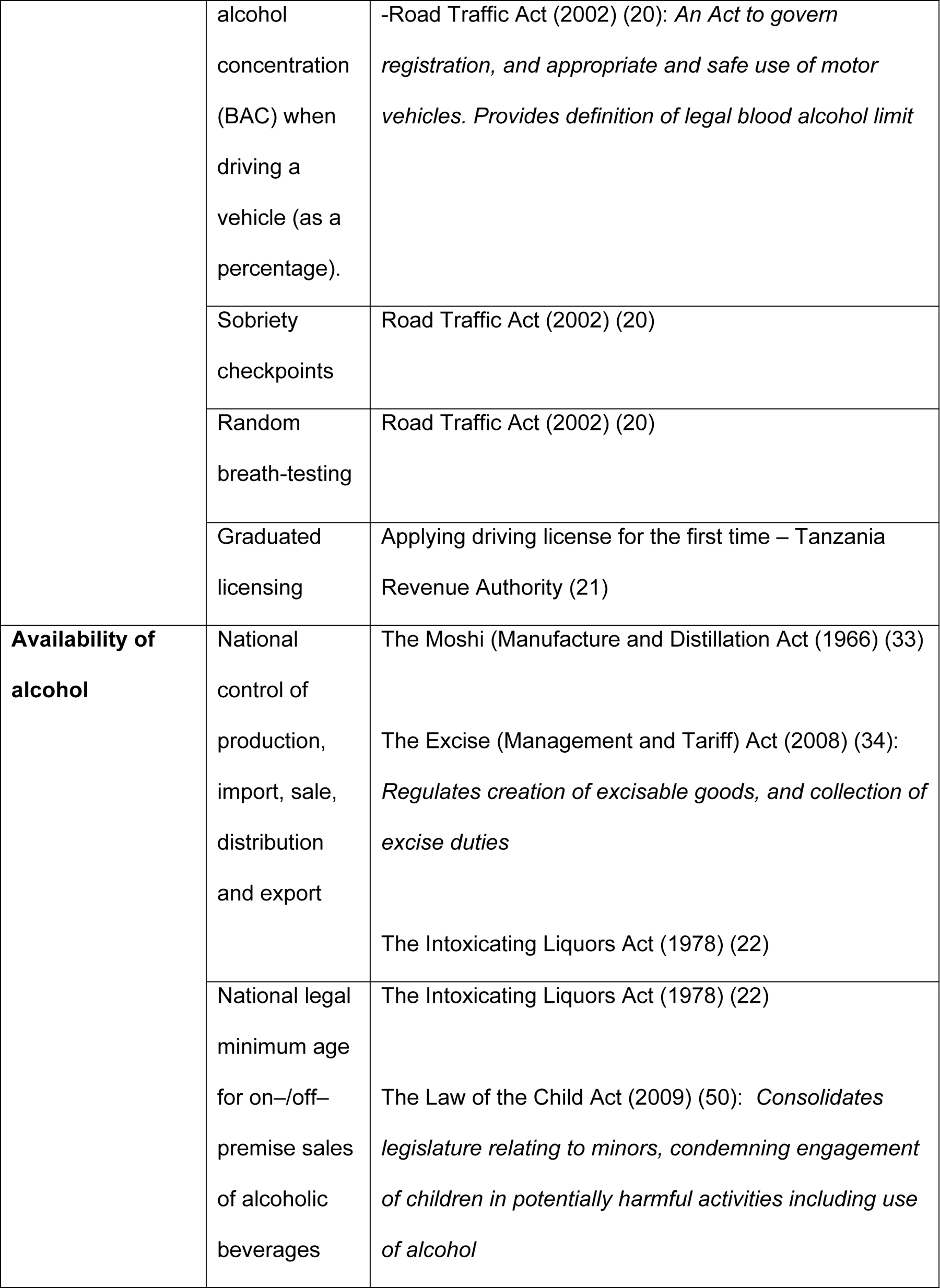
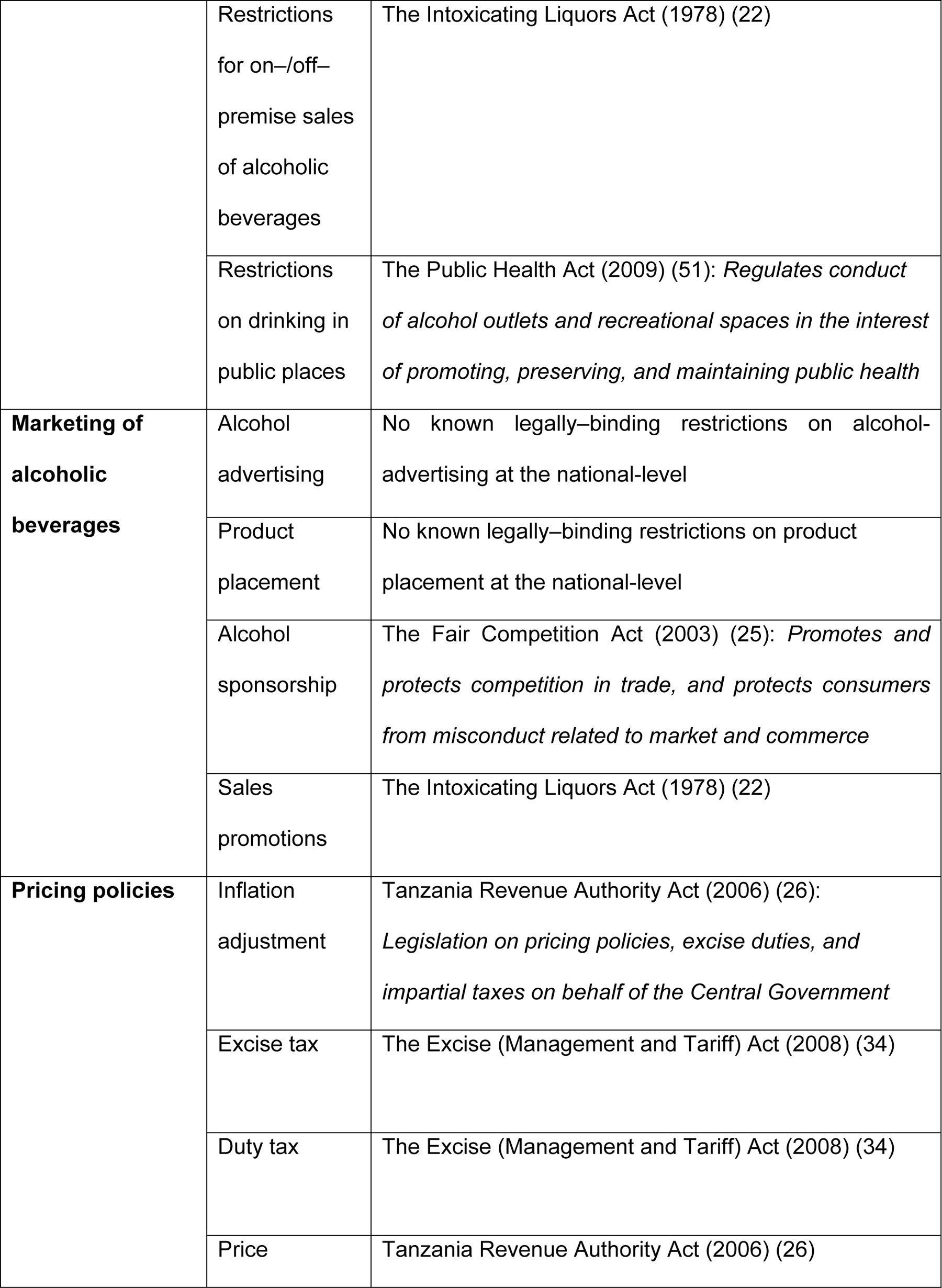
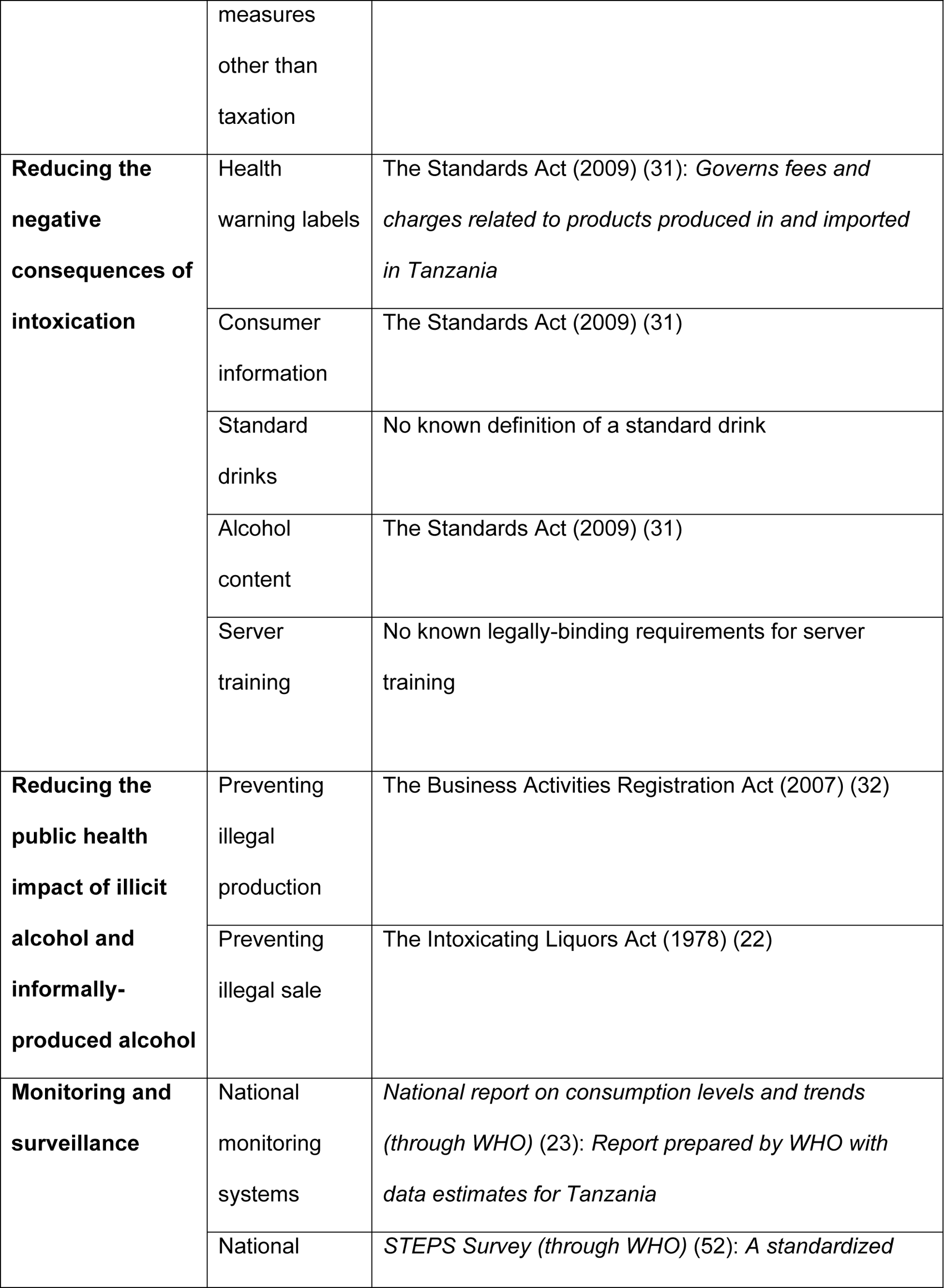
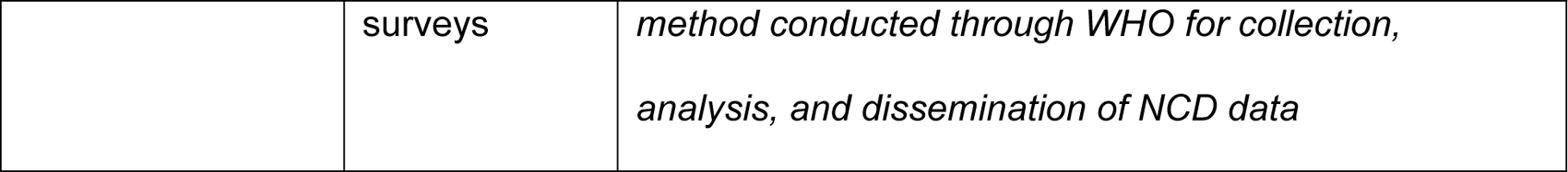
Relevant policy-related documents Error! Reference source not found.

### Leadership, awareness and commitment

The WHO framework states that member states should exhibit leadership and create awareness around challenges associated with alcohol consumption within their respective countries. They should also demonstrate commitment to controlling such challenges through adequately funded, comprehensive and intersectoral national policies (2).

Our searches and IDIs revealed no national alcohol policy or subsequent action plan.

> **Participant #5**: “First, we do not have such a [alcohol] policy so far. A national alcohol policy - which was being developed through the bench where I am, but it is a draft. I think - to be made about five to seven years ago. I think it lacked financing”
>
> **Participant #3**: “I don’t think if there is a policy that talks about the use of alcohol”

Pending a single all-inclusive policy, various documents and government authorities govern activities related to alcohol. There is little intersectoral linkage between these documents and bodies.

> **Participant #5**: “There is a need for cooperation in the various enforcement agencies so that so that the implementation can be done well”

Imposing further restrictions on the alcohol industry and committing to a national policy was consistently reported as a low priority by respondents.

> **Participant #5**: “There is the EAC regional policy [that talks about preventing, controlling and managing alcohol and drugs in the EAC]. But if it has guidance, if it affects laws, if it has an effect here in Tanzania, I haven’t seen it yet”
>
> **Participant #3**: “We have participated in the movement to prepare the National Youth Council. The law was passed five or six years ago but the Council has never been founded. If the council is formed, it will develop youth throughout the country. That is where we talk about violence, alcohol, and other deviant behaviour”

Overall, participants’ responses indicated that the development of a national alcohol policy is a low priority, particularly because alcohol is a lucrative source of revenue.

> **Participant #1**: “I don’t think [it is a priority]. Because we all know very well through alcohol, they get revenue”
>
> **Participant #2**: “I have not seen it [alcohol policy] as something that has been given much priority. Because alcohol is everyone’s favorite but also… it is the country’s economy”
>
> **Participant #8**: “I don’t think it is a priority”
>
> **Participant #6:** “So it [alcohol policy] is that the issue is not prioritized? Of course”

The government creates and encourages awareness activities on alcohol, often under the category of Non communicable diseases (NCDs) (12). This is done primarily through Ministry for Education, Science, and Technology (MoEST), and the Ministry of Health (MoH). Such awareness campaigns often highlight alcohol as a risk for NCDs. The Tanzania government recently launched the NCD Research Agenda and NCD Strategic Plan version 3 to guide these efforts (13). However, community-based organizations have also played a key role in motivating the government to create awareness and commit to this challenge, as reported by a respondent:

> **Participant #3**: “They [several Ministers] saw the importance of my organization to start meeting with students to motivate them and remind them about the contents of the policies that concern them. Those students have not found platforms”

### Health services’ response

Member states are required to use a multi-faceted and multi-level approach to create effective alcohol policies; individual-, family-, and community-level interventions are necessary through the health sector as a key entry point for people and families who are at risk of harmful alcohol use. The health sector is well-placed to leverage screening and awareness programs, treatment services, and advocate for community support (2).

Our searches identified the NCD Program as responsible for monitoring and reporting issues related to NCDs including health conditions caused by and related to alcohol use. However, there are no known focal national-level individuals or organizations allocated to respond specifically to alcohol issues alone.

> **Participant #3**: “In terms of alcohol, there is no board that has been created by the government”

Recent literature has shown a missed opportunity for screening at the primary-care level (14). While brief interventions, both psychosocial (education for individuals and families, motivational interviews, referral to support groups) and pharmacological are performed in out- and in-patient settings, these are often unaffordable, and there are no standalone treatment guidelines for alcohol-related issues.

> **Participant #2**: “I feel that they do not monitor the issue of alcohol. What they are following is to check his health, maybe with reproductive issues. You find it is difficult to monitor because it is not the main activity there”
>
> **Participant #6**: “I can say it is challenging. Many hospitals still do not have psychological services. 70%, they cannot afford. There are others who have to be referred to a hospital [for inpatient care] like Lutindi [more than 300km away from here]”

Overall therefore, insufficiency of mental health professionals and low integration of mental health in primary healthcare (15), means that screening for alcohol use is almost exclusively done within specialty-care settings, which is often less affordable and accessible (16). Likewise, the available yet insufficient rehabilitation institutions often go beyond their designated scope of work to compensate for an increasing demand.

### Community and workplace action

WHO recommends that member states and other implementers support and encourage communities to be proactive in reducing harmful alcohol use. WHO further recommends performing rapid assessments to identify priorities, and disparities in knowledge and practice within these communities (2).

The Tanzanian government has made some commitment to supporting community action, with limited sustainability. One move is attempting to standardize care for people struggling with use of alcohol and other substances by formulating national guidelines for all sober houses in the country, finalized in 2018. According to these guidelines sober houses should provide supportive and controlled living conditions, as well as psychosocial interventions for people with substance use (and often other related mental health) challenges (17). Due to insufficient rehabilitation programs in Tanzania, sober houses often provide both rehabilitation and transitional services as individuals prepare to re-enter their communities (18).

#### • School-based action

While MoEST has created awareness campaigns against harmful use of substances including alcohol, these happen infrequently, and are particularly rare at the university level, where most students are of legal consumption age. There is a lack of youth awareness and prevention programs, and the necessity for youth engagement in community action.

> **Participant #7**: “The issue of alcohol is included very little on the curriculum. If we go to colleges, they are very specialized and very rare to teach about alcoholism. Maybe it will happen when we discuss another topic, for example, “If you want to succeed in your career as a professional, things that you need to avoid [include] alcoholism”. But there is no topic or sub-topic or course or program”
>
> **Participant #7**: “I’m sure I can’t name a page or chapter that lets them know the harm of alcohol. The [Education] policy insists we make school-leavers or university-leavers who can use education to master the environment and their lives. It emphasizes that but not directly”

The Tanzania Commission for Universities stipulates higher education institutions create conducive environments for proper conduct among students and staff. Universities then create regulations which serve as technical tools to guide disciplinary action and healthcare assistance for students who present with behavioral changes, including those due to alcohol.

> **Participant #7**: “Regulations are created by colleges, but Tanzania Commission for Universities also points out it is appropriate for each institution to have examination regulations and those other disciplinary guidelines”

In an attempt to fill this support gap, various non-governmental organizations (NGOs) offer community-based interventions in Tanzania; Alcoholics Anonymous (AA) is the most widespread CBO across Tanzania, with seven active support groups in different cities (19). Other NGOs have created awareness on alcohol and linked people to care, but frequently as part of a program for Human Immunodeficiency Virus (HIV) care (11).

> **Participant #1**: “They [government officials] were very happy and said they didn’t know if there is such a thing (Alcoholics Anonymous) here in town, they wanted us to give them our contacts. It’s rare to get an invitation directly from the government. I haven’t seen them again”

### Drink-driving policies and countermeasures

WHO recommends establishing legal limits on blood-alcohol content while driving, the use of sobriety checkpoints, and sanctions for infraction of these regulations. It is further recommended that drivers’ licenses be provided on a multiple phase system to ensure sufficient training (2).

Tanzania has defined a maximum prescribed blood-alcohol limit for drivers, and requires enforcement of checkpoints for selective blood- and breath-testing for suspected driving under influence of alcohol or actual hazardous driving, and penalties for determined offences (20). A graduated licensing system is currently in place (21). Interviewees consistently highlighted poor integration of enforcement agencies, prioritization of driving offences other than alcohol-related ones, and infrequent utilization of sobriety checkpoints.

> **Participant #2**: “They [traffic police] will look at other mistakes, “you have overtaken, speeding, you haven’t paid insurance”, but they don’t say “Let’s stop him and check if he is drunk.”
>
> **Participant #5:** “The willingness of those enforcement agencies to act… maybe they haven’t had direction. It requires relations with other governmental and legal institutions, to be able to show the difference”

### Availability of alcohol

WHO recommends restrictions on production, retailing and distribution, legal age of purchase, and drinking in public spaces as means to achieve this (2).

Various documents comment on regulating availability of alcohol in Tanzania; restrictions on sale of alcohol by time and place are provided, as well as governance on handling of already intoxicated persons. However, there are no restrictions for public environments such as petrol stations or specific events including weddings and concerts (22,23). While an age limit of 18 years is stated for purchase of alcohol both on- and off-premises, interviewees frequently reported traditions from across Tanzania that allow and encourage children to engage in alcohol production, distribution, and consumption, particularly in Northern Tanzania.

> **Participant #1**: “At home alcohol was used in every event, recreation, celebration. Boys starting from twelve or fourteen [age] serve elderly men. He asks you to taste a little then you give it to him. It was regarded as culture. It’s normal”
>
> **Participant #4**: “Many have started [using alcohol] at a younger age”
>
> Participants noted that alcohol is highly profitable, and retailers may prioritize income over safety of underage customers.
>
> **Participant #2**: “Sometimes people don’t think about safety of those young people, what they look at is money, how much they earn – “If I stop selling to children, they will run elsewhere, and I may be fired because I have chased away customers” **Participant #6**: “Under 18, it’s not allowed. The law exists but people do not enforce it”
>
> Another interviewee highlighted the practice of unregulated re-packaging and sale of alcohol in public spaces. This is in contravention with the ban on alcohol sachets (24), due to their affordability and inconspicuous packaging, therefore, also easily accessed by youth.
>
> **Participant #6**: “Strong alcohols being sold at bus stops. They sell them in small glasses ranging from 500, 1000 or 2000 shillings. They also sell to adolescents”

### Marketing of alcoholic beverages

WHO’s global strategy recommends the use of legally-binding restrictions on marketing, sponsorships, promotions, and product placement that may directly or indirectly target youth; this includes modern marketing strategies such as social media (2).

One of our respondents indicated that alcohol advertisements are often self-regulated by media outlets, however, decisions are heavily influenced by peak engagement times.

> **Participant #8**: “These media houses, all they think is money. So, it depends on what time of day they can get the most listeners. For example, on [name of radio program redacted for confidentiality purposes] in the morning, we’ve had ads [on alcohol] go up”

There are partial restrictions on sponsorship of events; local brands are prioritized over foreign ones, particularly for sporting events. Alcohol companies are also not allowed to be the sole sponsor of an event as guided by regulations on fair competition (25).

> **Participant #8**: “I don’t think they are allowed to sponsor everything. They give a certain percentage of money to invest for like, a marathon, concerts, or football. And to mostly be a Tanzanian brand”

There are also partial restrictions on sales promotions; auctioning of alcohol products is permissible with appropriate license and premises (22). However, the Act does not comment on free- or below-cost promotions by retailers. Furthermore, our searches and interviews did not produce results on national-level legally-binding regulations on product placement.

### Pricing policies

WHO requires member states to establish a system for adjusting prices of alcohol beverages to account for inflation, excise and duty taxes, with effective implementation and review mechanisms (2).

The Tanzania Revenue Authority (TRA) oversees pricing policies in the country; to instate, evaluate, collect, and adjust excise and duty-paid taxes on industrial alcohol products, thereby controlling prices. TRA enforces laws related to these taxes (26,27). However, estimates show that informally produced alcohol, which is often unregulated, constitutes 86% of all alcohol consumed in Tanzania (23), indicating that TRA has little control over majority of alcohol producers.

> **Participant #5**: “The willingness of those enforcement agencies to act, there may be differences”

Reports reveal the alcohol industry as a growing source of revenue for the government (28). Locally-produced alcohol is also relatively more affordable and easily accessible than industrial alcohol. Furthermore, current pricing policies do not account for disproportionate income levels across different parts of Tanzania (5), and most commonly consumed alcohol products remain affordable even for those without reliable income (29).

> **Participant #6**: “These strong alcohols are still very cheap; it becomes a great influence especially for young people”

### Reducing the negative consequences of intoxication

To mitigate negative consequences associated with drinking alcohol, policies should take a harm-reduction approach; to prevent harm among those most at risk. WHO recommends establishing and enforcing legislation against serving already intoxicated customers, ensuring sufficient server training, and requiring informative labels about alcohol-related harm (2).

Tanzania legally requires health warning labels on alcoholic drinks to promote awareness on age restrictions and drinking in moderation. The Tanzania Bureau of Standards has regulated these inclusions to standardize alcoholic products, as well as other items produced in Tanzania (30).

> **Participant #8**: “They include a tagline “you can’t sell alcohol to someone who is below 18 years”

Furthermore, alcohol manufactured industrially must display alcohol content and ingredients on containers (31), however, there are no requirements for information on contents or additives on labels.

In connection to the lack of a definition for a standard drink, there are no legal requirements for displaying number of standard drinks on retail containers. Tanzania also lacks legally-binding requirements for training of persons involved in distribution of alcohol products; recruitment of drink servers and sellers is unregulated (23).

### Reducing the public health impact of illicit alcohol and informally-produced alcohol

WHO recommends developing monitoring systems and imposing the same regulations on informally-produced or unrecorded alcohol as with industrial alcohol. Member states are required to develop monitoring systems for the detection of such alcohol, to issue warnings on the health risks associated with their consumption, and to impose the same regulations on unrecorded alcohol as with industrial alcohol (2).

Tanzania has numerous policy-related documents in place that legally require registration of alcohol producers (32), regulation of manufacturing (33), and retail and distribution of alcohol (22,34). However, since more than 80% of consumed alcohol in Tanzania originates from informal producers (23), coupled with low enforcement, the impact of these regulations is likely to be low, and particularly in rural areas.

### Monitoring and surveillance

WHO recommends national surveys on patterns of alcohol consumption and related harms, and establishment of bodies to supervise research and reporting of findings (2).

All IDIs and online searches indicated lack of national-level surveillance of alcohol consumption patterns, health and social consequences, and alcohol policy responses.

WHO has reported data on alcohol consumption levels and trends in Tanzania as part of a larger international survey (23). The STEPwise approach to NCD risk factor Surveillance (STEPS) Survey, also conducted on an international level through WHO, last reported on determinants of NCDs in Tanzania including alcohol in 2012 (35). Furthermore, there has been a rise in research activities investigating determinants and outcomes of alcohol consumption in Tanzania over the past decade (5,11,36,37).

## Discussion

This review has revealed valuable insights into policies, laws, acts, and actions regarding alcohol use in Tanzania. There is no national alcohol policy in Tanzania. By using the framework of composite indicators created by WHO, we explored the extent to which alcohol policy is being enforced in the country. Our results indicate a lack of engagement between key sectors, and enforcement, which can have a detrimental effect on effectiveness of existing regulations, and make alcohol more available; this phenomenon has been observed in a previous policy analysis (38). A regional policy scoring scheme was conducted in 46 African countries and revealed Tanzania in 22^nd^ place, with medium restrictiveness. This study found a significant association between lower restrictiveness and higher per capita alcohol consumption (39).

While the EAC regional policy document guides prevention programs, treatments services and coordination of policy, its framework is limited in comparison to the WHO Global Strategy; the former has fewer indicators of implementation, and is derived from and influenced by literature on HIV, illicit drugs as well as alcohol. These key differences guided our selection of the WHO document as our reference for the study.

Lack of financing has partly hindered finalization of the national alcohol policy draft. There is minimal intersectoral linkage between various existing regulations and enforcement agencies; minimal to no regulation on marketing of alcohol, pricing policies, and national-level surveillance of consumption trends and socio-economic and health outcomes. Commitment from the government is more impactful on industrial alcohol producers, which accounts for less than 20% of consumed alcohol in the country (5). Furthermore, the majority of our findings indicate that alcohol policy is a relatively low priority for the government, and that an existing regional policy has negligible impact on national-level implementation.

### Interpretation and integration with previous literature

Our research adds to the limited previous literature in this area. For example, a 2012 situational analysis highlighting resources across Tanzania related to alcohol use and its regulation, focused primarily on HIV prevention (11). By contrast, our use of the WHO framework allowed a more comprehensive assessment and comparability with other countries. WHO provides clear guidance on requirements for its member states; Firstly, a primary responsibility for formulating, implementing, monitoring and evaluating public policies to reduce the harmful use of alcohol. The lack of integration and engagement noted in our findings is to be expected given the lack of a single comprehensive policy. This is similarly emphasized in a regional assessment of alcohol policy in African countries, whereby re-evaluation and increasing restrictiveness and integration of policy was recommended (39). Our findings also suggest that through utilizing a socioecological approach, the health sector can support multi-level interventions and advocate for support within communities at large. This is supported by previous literature from Tanzania on missed opportunities to addressing motivators and barriers to seeking care (14).

Stricter regulations on physical access to alcohol are an effective, sustainable, and feasible approach to counteract inappropriate or harmful alcohol use; this is particularly important amongst the young. Marketing restrictions, by extension, are a key point of intervention to minimize early and inappropriate exposure to alcohol advertising. Our findings indicated strong socio-cultural drivers for underage drinking, therefore, strategies to address such practices are paramount. Prospective literature and recent studies from Tanzania similarly indicate that exposure to outdoor advertisements is one of the primary reasons for alcohol initiation among youth (37,40), and high-density advertising facilitates underage drinking (5). The Tanzanian government has recently made an effort to minimize physical access to alcohol among youth by banning the more affordable alcohol sachets (41). However, our findings indicate that street vendors have found ways to circumvent these restrictions.

Raising the cost of alcoholic beverages is one of the most effective strategies for minimizing hazardous alcohol use (42). Our findings revealed that prices of alcoholic beverages remain affordable to most consumers across all ages. Furthermore, unregulated re-packaged alcohol products are available to the public, and are easily accessed by those at risk. Previous literature has explored this gap in policy, and identified pricing regulations as successful in reducing rates of morbidity and mortality (43), violence (44), and unplanned underage pregnancies associated with alcohol use (45).

Illegally-imported and informally-produced alcohol (homemade and small-scale production beverages) represent unique subsets of consumed alcohol; their alcohol content may be more difficult to quantify, unknown ingredients may be included, they often cost less than industrial alcohol products, and levels of their consumption are difficult to monitor. Altogether this poses a challenge to member states’ efforts in regulating alcohol consumption. Lastly, monitoring and surveillance activities inform enforcement and implementation strategies and allow for more robust policies. Previous studies echo our findings, emphasizing the need for regulation of informal alcohol producers (46) and strengthening monitoring systems (47).

### Strengths and Limitations

To our knowledge, this is the first comprehensive mixed methods alcohol policy review in Tanzania. Additional incorporation of the WHO framework to guide data collection and structuring IDIs adds validity to our findings. It contributes to the previous scholarship on alcohol use in Tanzania, by systematically identifying gaps and opportunities in the alcohol policy landscape in Tanzania through the use of a pre-existing globally recognized framework for measuring policy implementation. Furthermore, by utilizing the WHO framework intended for international audiences, our findings by extension may also offer valuable insights for policy implementation gaps in other countries.

We aimed to maximize validity through the rigorous use of multiple methods including a thorough search process, and wide range of IDI participants ranging from primary healthcare delivery, curriculum development, marketing, research, policymaking, CBOs, and Ministry-level officials. These respondents represent settings where mental health and rehabilitation resources for people with a history of hazardous alcohol use are sparse; where people are exposed to production, distribution and advertisements of alcohol from a young age; are known to have little awareness of alcohol and its effects either through education or community programs; and where policy development work has faltered.

However, this study had several limitations. The majority of our respondents were representative of regional work and of urban settings, due to challenges in securing interviews with national-level representatives. These findings, therefore, may not provide an entirely accurate representation of enforcement across the country. However, since regulation of alcohol predominantly occurs in urban settings, we are confident in the applicability of these results within that context. Furthermore, several documents were excluded due to their inclusion under HIV programs; frequently, alcohol is discussed as a risk factor for HIV and NCDs. Yet, little has been done to address socio-cultural complexities around alcohol when included in this way. Pending its own policy and comprehensive implementation work, alcohol will likely remain in the shadows of other priorities. Undeniably, interviews with individuals regarding aspects of their own professional work may be subject to social desirability biases. However, our desk-review findings corroborate key opinions stated in the IDIs.

## Conclusion and recommendations

We have identified lack of overall policy development and co-ordination, poor integration, enforcement and regulation in relation to Alcohol policy in Tanzania. Our findings strongly suggest the need for a comprehensive approach to developing an alcohol policy, with key stakeholders and youth involvement, stronger enforcement, and building on awareness and resources for those affected by and at risk of harmful alcohol use, to address this issue of concern.

The health sector has the potential to be a key supporter of awareness activities, mobilizing research around alcohol consumption and its associated challenges, which in turn should inform policy. Further nationwide surveys and other specific studies could add depth to the understanding of alcohol-related harm in Tanzania. The presence and momentum of CBOs and modern marketing strategies are yet another opportunity for low- and middle-income countries such as Tanzania to collaboratively reach, support, and empower people in need. We strongly recommend increasing government support and collaboration with CBOs to improve knowledge on alcohol and standardization of services. Lastly, there is need to publicize and consistently utilize regulations which are already available.

## Data Availability

Deidentified datasets used and/or analysed during the current study are available through URL from the corresponding author on reasonable request.

## List of abbreviations

AA: Alcoholics Anonymous
CBO: Community-based organization
EAC: East African Community
HIV: Human Immunodeficiency Virus
IDI: In-Depth Interview
MoEST: Ministry of Education, Science and Technology
MoH: Ministry of Health
NCD: Non-Communicate Diseases
NGO: Non-governmental organization
SSA: Sub-Saharan Africa
TRA: Tanzania Revenue Authority
WHO: World Health Organization

## Acknowledgements

Sincere gratitude goes out to the respondents for their time and invaluable experience.

## Declarations

### Consent for publication

Not applicable

### Authors’ contributions

K.M. led on methodology, study co-ordination, writing, preparation of tables and figures, and review & editing. A.H. contributed to methodology, writing, review & editing. M.L.S. contributed to project administration, methodology, study co-ordination, writing, and review & editing. A.O. and B.M. contributed to review & editing.

### Patient and Public Involvement

Patients or the public were involved in the design, or conduct, or reporting, or dissemination plans of our research.

